# Data-driven modeling and forecasting of COVID-19 outbreak for public policy making

**DOI:** 10.1101/2020.07.30.20165555

**Authors:** A. Hasan, E.R.M. Putri, H. Susanto, N. Nuraini

**Affiliations:** Mærsk McKinney Møller Institute, University of Southern Denmark, Denmark; Department of Mathematics, Institut Teknologi Sepuluh Nopember, Indonesia; Department of Mathematics, Khalifa University, United Arab Emirates; Department of Mathematical Sciences, University of Essex, United Kingdom; Department of Mathematics, Institut Teknologi Bandung, Indonesia

**Keywords:** COVID-19, modeling, forecasting, public policy

## Abstract

This paper presents a data-driven approach for COVID-19 outbreak modeling and forecasting, which can be used by public policy and decision makers to control the outbreak through Non-Pharmaceutical Interventions (NPI). First, we apply an extended Kalman filter (EKF) to a discrete-time stochastic augmented compartmental model to estimate the time-varying effective reproduction number ℛ_*t*_. We use daily confirmed cases, active cases, recovered cases, deceased cases, Case-Fatality-Rate (CFR), and infectious time as inputs for the model. Furthermore, we define a Transmission Index (TI) as a ratio between the instantaneous and the maximum value of the effective reproduction number. The value of TI shows the disease transmission in a contact between a susceptible and an infectious individual due to current measures such as physical distancing and lock-down relative to a normal condition. Based on the value of TI, we forecast different scenarios to see the effect of relaxing and tightening public measures. Case studies in three countries are provided to show the practicability of our approach.

## 1. Introduction

The spread of new coronavirus disease 2019 (COVID-19), originating from Wuhan China, has been worldwide and caused a severe outbreak. The virus has infected more than 17.5 million people with more than 680 thousand confirmed deaths by the end of July 2020 [1]. The outbreak triggered crisis which is beyond health crisis and it is expanding into economic and social crisis. The effect of crisis to the society become a multidimensional problem that need to be minimized through some measurable public policies [2].

### 1.1. Motivation

Intervention measures are introduced to contain the outbreak and to prevent it continuing to grow and transmitted, such as physical distancing and lock-down measures [3, 4]. To this extent, a thorough evaluation to implement available options, has been urgently needed. A quantitative as well as a qualitative evaluations involving key characteristics of COVID-19 outbreak can be conducted based on epidemiological parameters [5]. As the incidence is growing, a quantitative evaluation to identify a minimum physical distancing policy aims to control the outbreak transmission in Australia [6], China [7], and Italy [8].

A control measure for the disease’s transmission, known as the time-varying effective reproduction number ℛ_*t*_, reflects the disease extended transmission with the presence of interventions. Therefore, estimation of the time-varying effective reproduction number ℛ_*t*_ can be used for evaluating implementation of public policies success [9]. The estimation of ℛ_*t*_ based on an epidemiological model, has an important role for an evidence-based policy making. The importance is also recognized by the World Health Organization (WHO) [10].

### 1.2. Literature review

A deterministic SIR model-based for ℛ_*t*_ estimation, assumes that the data used significantly representing the actual outbreak, was presented in [11]. Different sets of data representing level of quarantine measures, are used in [7] describing growth of the cases and also the effective reproduction number ℛ_*t*_ for each measure’s levels. To accommodate uncertainties in incidence data, noise is added to the model in [9]. The authors used inputs for the model from daily new cases, active cases, recovered cases, and deceased cases, to estimate the spread of the disease and the evaluation is extended to ℛ_*t*_ estimation based on the stochastic model.

Based on estimation of ℛ_*t*_, several authors have proposed methods to forecast the evolution of the outbreak. Data-based analysis, modelling, and forecasting based on a Susceptible-Infectious-Recovered-Deceased (SIRD) model was presented in [12]. The authors fit the reported data with the SIRD model to estimate the epidemiological parameters. The main drawback when fitting the model with the data is that the estimated parameters can be unrealistic. In [13], the authors attempted to use phenomenological models that have been validated during previous outbreaks. The model is used to generate and assess short-term forecasts of the cumulative number of confirmed reported cases. However, since COVID-19 is a new virus, the model was not reliable and the forecast can only be used for a very short term. Another authors use analysis of simple day-lag maps to investigate universality in the epidemic spreading [14]. Their results suggested that simple mean-field models can be used to gather a quantitative picture of the epidemic spreading. The main drawback is that the reproduction number is assumed to follow a heuristic continuous model, which may not describe the actual transmission.

### 1.3. Contribution of this paper

In this paper, we propose a data-driven approach for COVID-19 modeling and forecasting, which can be used by public policy and decision makers to control the outbreak through Non-Pharmaceutical Interventions (NPI). Considering drawbacks in existing methods, we present two contributions: (i) estimation of the time-varying effective reproduction number ℛ_*t*_ based on real-time data fitting using an extended Kalman filter (EKF), and (ii) short to medium terms forecasting based on different public policies. As the effective reproduction number ℛ_*t*_ shows simply the extent of transmission due to population immunity or intervention in the form of public policy making [9], we propose a new measure called a Transmission Index (TI) (see Section 2.5), which describe the disease transmission relative to a normal condition. As well as ℛ_*t*_, the value of TI can be used to measure the effectiveness of public health measures. Furthermore, TI is used to forecast different public policy scenarios by relaxing or tightening the current measures.

### 1.4. Organization of this paper

Briefly, this paper is organized as follows. In Section 2, we discuss the methods used in evaluating the spread of the disease, including data availability and reliability, data driven framework, modelling, estimation, and forecasting. Then, discussion about the method and its applications for estimating the Transmission Index in United Arab Emirate (UAE), Australia, and Denmark, is presented in Section 3. Lastly, the conclusion is in Section 4.

## 2. Methods

In this section, we describe the data-driven modeling and forecasting approach that can be used by public policy and decision makers to control COVID-19 pandemic through NPI. We acknowledge no country knows the total number of people infected with COVID-19, partially due to lack of testing and undetected asymptomatic cases. Thus, the presented approach can only be used for country/region/area that have performed mass testing with laboratory confirmation. To this end, we assume the difference between the actual case and the reported case is minimized when mass testing has been conducted.

### 2.1. Data availability and reliability

The COVID-19 pandemic generates a large amount of data. Typically, the government officials reported daily confirmed cases, active cases, recovered cases, and deceased cases (see Table 1). These data are available for almost all countries and regions and can be accessed by the public through online websites. Some websites, such as https://www.worldometers.info and https://ourworldindata.org/, also provide information regarding the number of test per capita. The data can be utilized to obtain important epidemiological parameters such as the time-varying effective reproduction numbers ℛ_*t*_, which can be used by the decision and public policy makers.

**Table 1:** A typical time-series data generated during the pandemic.

| Date | Month | Daily Confirmed (C) | Active Case (I) | Recovered (R) | Deceased (D) |
| --- | --- | --- | --- | --- | --- |
| ⋮ | ⋮ | ⋮ | ⋮ | ⋮ | ⋮ |
| 28 | 4 | 25498 | 830745 | 148926 | 59418 |
| 29 | 4 | 28525 | 851065 | 154737 | 61812 |
| 30 | 4 | 30912 | 874215 | 160293 | 64018 |
| 1 | 5 | 36090 | 898527 | 170171 | 65918 |
| 2 | 5 | 29816 | 914246 | 182570 | 67616 |
| 3 | 5 | 27394 | 934901 | 188155 | 68770 |
| ⋮ | ⋮ | ⋮ | ⋮ | ⋮ | ⋮ |

Unfortunately, not all countries/regions have the ability to provide mass testing for their citizen. The WHO advised governments that the positivity rate (i.e., out of all tests conducted, how many came back positive for COVID-19) should remain at 5% or lower for at least 14 days [1]. Rich countries such as the UAE, Australia, and Denmark have successfully achieved this target.

### 2.2. Data-driven framework

The reported cases from the pandemic are used for two purposes: (i) to estimate the time-varying effective reproduction number ℛ_*t*_, and (ii) to project the number of active case, recovered case, deceased case, and total case, which are important to prepare for the healthcare systems.

In Figure 1, *S* denotes susceptible case data. Assuming constant population, *S* can be obtained by subtracting the number of population with the number of active, recovered, and deceased cases. The active case data *I* is the number of people who are currently infected. *R* and *D* denote the cumulative number of recovered and deceased, respectively, while *C* denotes the number of daily confirmed cases. To model uncertainty in the reported cases, we add white Gaussian noise. The data is assimilated into a compartmental epidemic model (see Section 2.3) using an extended Kalman filter (EKF) to obtain the estimate of ℛ_*t*_. Based on this estimate, we perform forecasting for the next 90 days with different scenarios.

**Figure 1:**
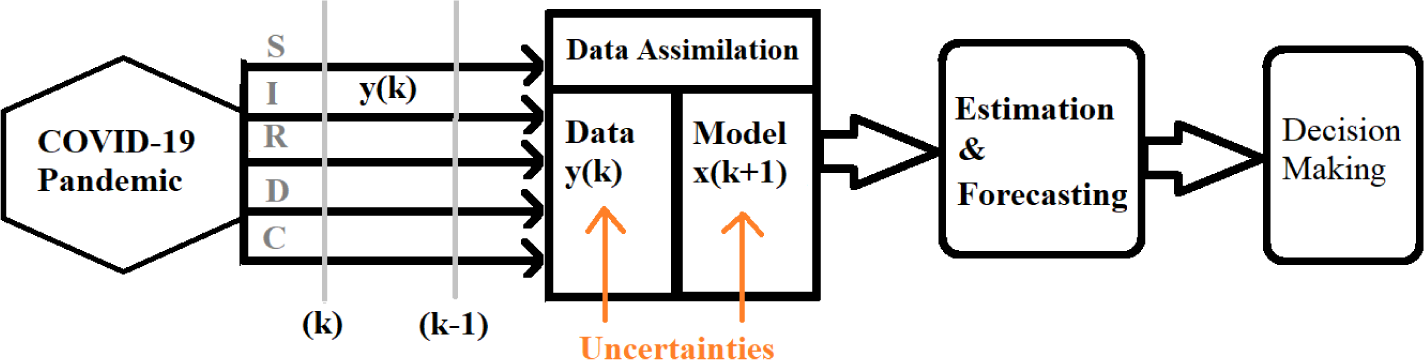
Data-driven framework of COVID-19 pandemic.

### 2.3. Mathematical modeling

To model the transmission of the coronavirus, we use a discrete-time stochastic augmented compartmental model presented in [9]. The model consists of six equations

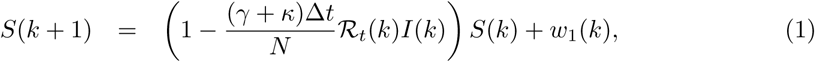

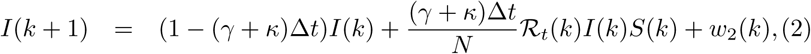

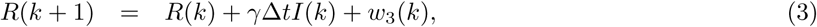

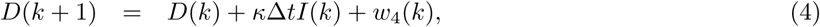

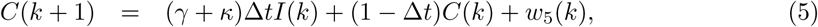

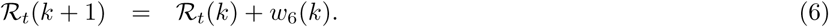

The equations (1)-(4) can be obtained from a standard Susceptible-Infectious-Recovered-Deceased (SIRD) model. We augment two equations: equation (5) takes into account the number of daily confirmed cases *C*, while equation (6) says that the effective reproduction number ℛ_*t*_ is assumed to be a piecewise constant function with jump every one day time interval. The noise *w*_1_(*k*), *w*_2_(*k*), *w*_3_(*k*), *w*_4_(*k*), *w*_5_(*k*), *w*_6_(*k*) are used to model the uncertainty.

The discrete-time stochastic augmented compartmental model (1)-(6) has three constant parameters: the number of population *N*, the recovery rate *γ*, and the death rate *κ*. The recovery and death rates are depend on infectious time *T*_*i*_ and Case-Fatality-Rate (CFR), and are given by

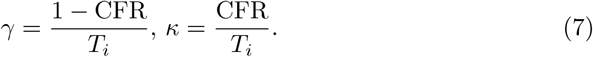

The infectious time is obtained from clinical data. For COVID-19, the infectious period on average lasts for 9 days with standard deviation of 3 days [15]. The CFR is unknown and need to be estimated. However, to simplify the calculation, in this paper we assume the CFR is equal to the last data of the number of deceased case divided by the total infected case. To account for under-reported case, this estimate can be divided by a correction factor *c*, e.g.,

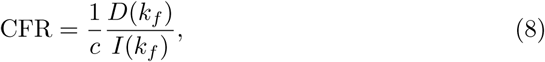

where *k*_*f*_ denotes the index for the latest data. In our example in Section 3, we assume the under-reported case is 3 times larger that the reported case. Thus, we take *c* = 4.

### 2.4. Estimation of ℛ_t_

The time-varying effective reproduction number ℛ_*t*_ is estimated by applying EKF to the discrete-time stochastic augmented compartmental model (1)-(6). The details regarding the implementation can be found in [9]. The algorithm has two tuning parameters: the covariance of the process noise ***Q*** and the covariance of the observation noise ***R***, which can be chosen such that the Root Mean Square Error (RMSE) between the reported and estimated data is minimized. The EKF serves as a real-time data fitting. The EKF will estimate any new data that is recorded. Once this estimation process works, the EKF will also produce an estimate of ℛ_*t*_ from (6).

Figure 2 shows results from implementing EKF to (1)-(6) using data from the United Arab Emirates (UAE). It can be observed that the EKF is able to estimate the active, recovered, deceased, and daily confirmed cases accurately. This data fitting process produces an estimate of ℛ_*t*_ based on correlation in the model (1)-(6).

**Figure 2:**
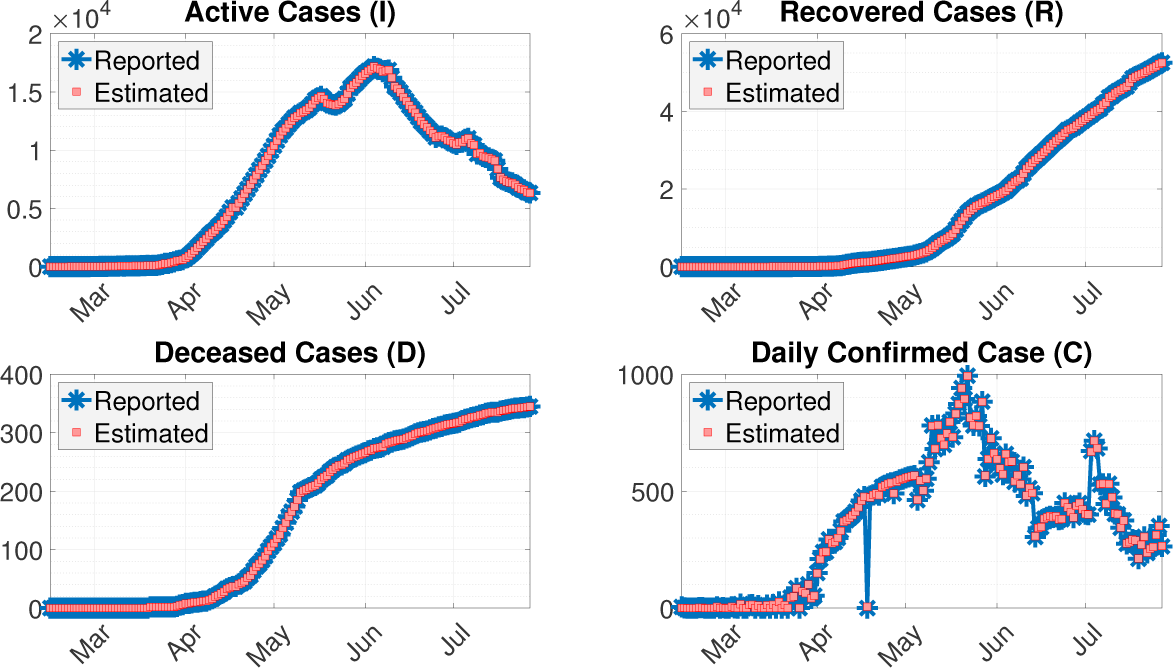
An example of real-time data fitting using EKF.

### 2.5. Forecasting

Forecasting are done for different scenarios. To this end, we define a Transmission Index (TI) as

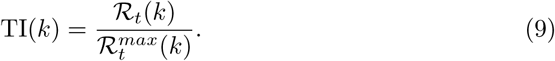

Figure 3 shows an example of different value of TI to simulate different public measure strategies. In this case, we draw a straight line perturbed by a white Gaussian noise between the current TI (at blue vertical line) with a designated TI (after 90 days). The noise is added to simulate fluctuation in the reported cases. The designated TI corresponds to actions to be taken by the decision or public policy makers. In this example, the designated TI are from 15% to 90%. The 30-day average of TI (plotted in black line) represents the value of TI due to current public measures. Here, the 30-day average of TI is at 51%. If the public measures are relaxing, the value of TI could go from 51% to 75% or 90%. Vice verse, if the public measures are tightening, the value of TI could go from 51% to 35% or 15%. Based on different value of TI, we use the discrete-time compartmental model (1)-(5) to forecast the outcome of different scenarios.

**Figure 3:**
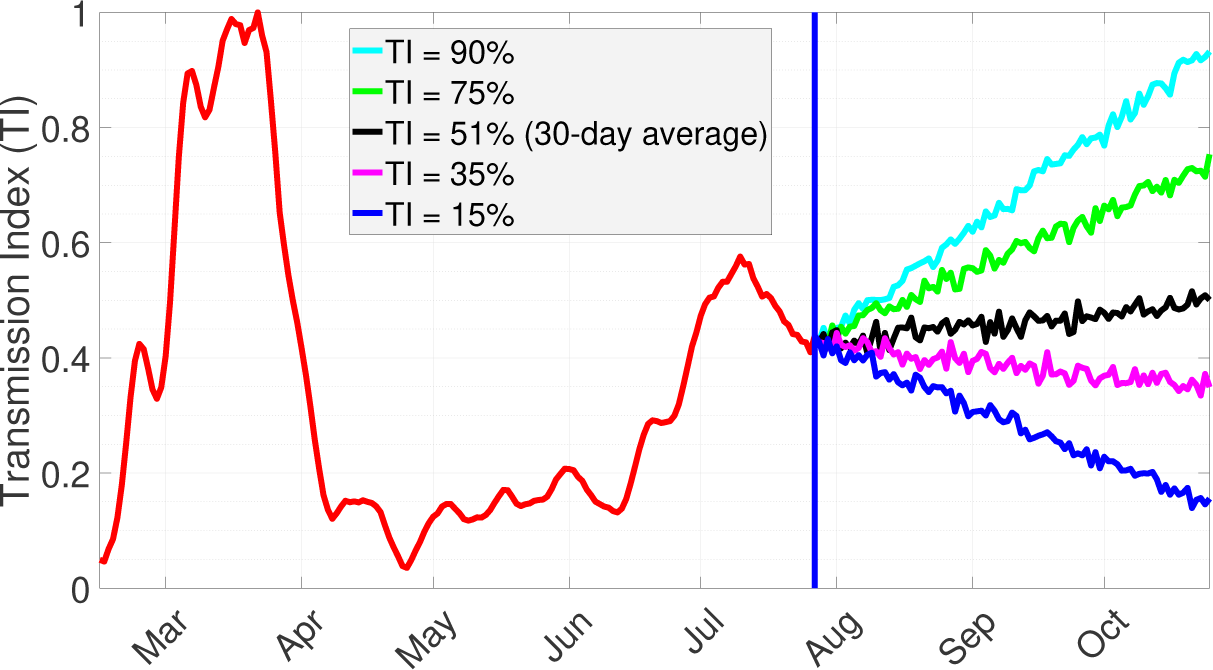
Forecasting for different scenarios are simulated using different designated value of TI.

## 3. Result and Discussion

In this section, we run simulations for three countries: UAE, Australia, and Denmark. All data sets and MATLAB codes are available on GitHub through this link: https://github.com/agusisma/coviddatadriven. Parameters for the simulations are presented in Table 2. The recovery rate *γ* and the death rate *κ* are calculated using (7).

**Table 2:** Parameter used in simulations.

| Country | Parameters |  |  |
| --- | --- | --- | --- |
| | N | CFR | $T_i$ |
| UAE | 9,890,402 | 0.15% | $9 \pm 3$ |
| Australia | 25,499,884 | 0.27% | $9 \pm 3$ |
| Denmark | 5,792,202 | 1.13% | $9 \pm 3$ |

### 3.1. United Arab Emirates

The UAE has conducted more than 4.9 million tests since the outbreak or 502.14 total tests per thousand population [16]. This brought UAE as one of the countries with the highest number of tests. The study of [17] estimates that the percentage of symptomatic COVID-19 cases reported in UAE using case fatality ratio estimates is at 98% (86%-100% of 95% credible interval).

The first confirmed cases were reported on 29 January, from an infected family of four who came to the country on holiday from Wuhan [18, 19]. As the number of positive cases steadily increased, the government took immediate public measures, such as the closure of schools and universities across the country until the end of the academic year in June (announced on 3 March) [20], the suspension of prayers at mosques and all other places of worship from 16 March including the whole month of Ramadan [21], as well as night curfews for disinfection on 26 March for an extended period of time that limited movements within the country [22].

The extreme measures together with the government’s wider National Screening Programme, which seeks to test as many people as possible with the aims to identify, isolate and treat patients as quickly as possible, yielded positive results in the decrease of the reproduction number almost immediately afterwards, see Figure 4. On 18 May the government announced the first day where the number of recoveries surpasses the number of new cases found [23]. In the third week of May, the daily new cases reached its peak and the reproduction number was already below the threshold value ℛ_*t*_ = 1.

**Figure 4:**
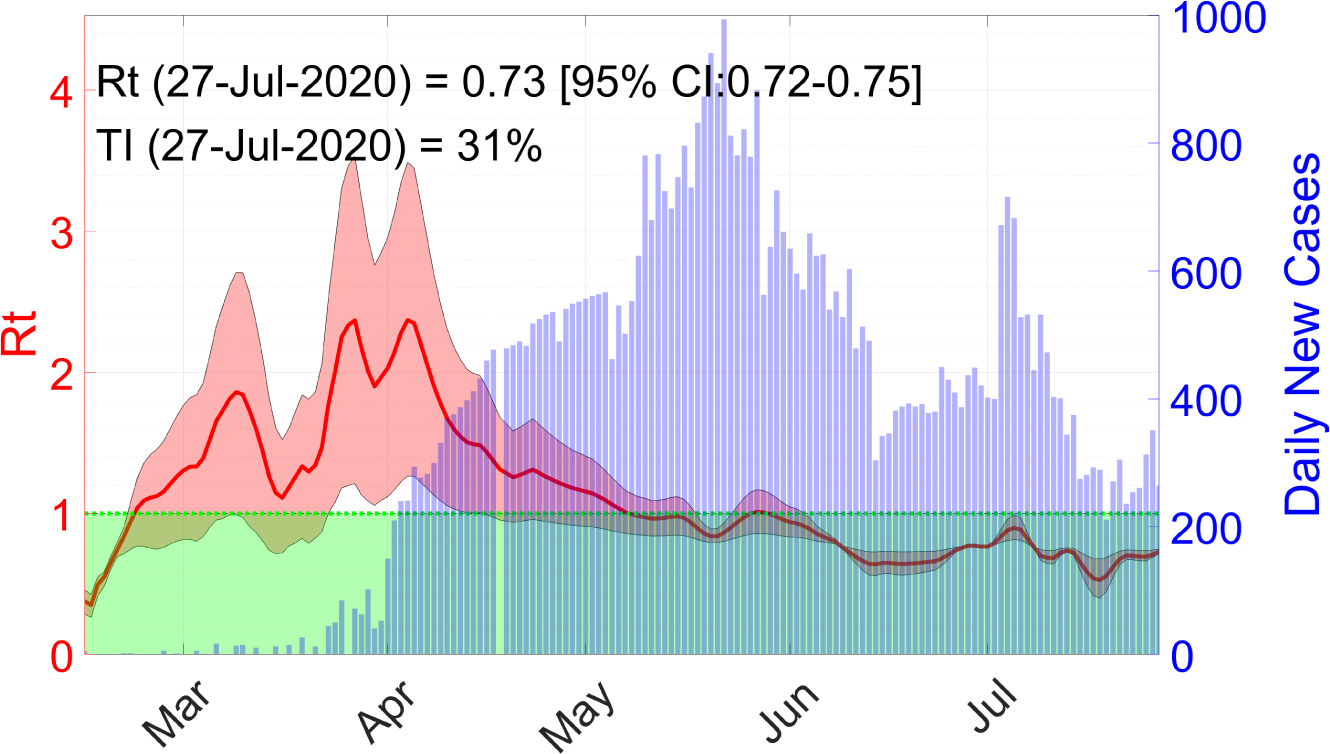
The time-varying effective reproduction number ℛ_*t*_ in UAE.

With the current TI at 31%, our forecast shows that the daily cases will be steadily decreasing, see Figure 5. As restrictions in the country ease, it is important to main safety and preventive measures. Public negligence can increase the TI, which can trigger a second wave of infection in the country.

**Figure 5:**
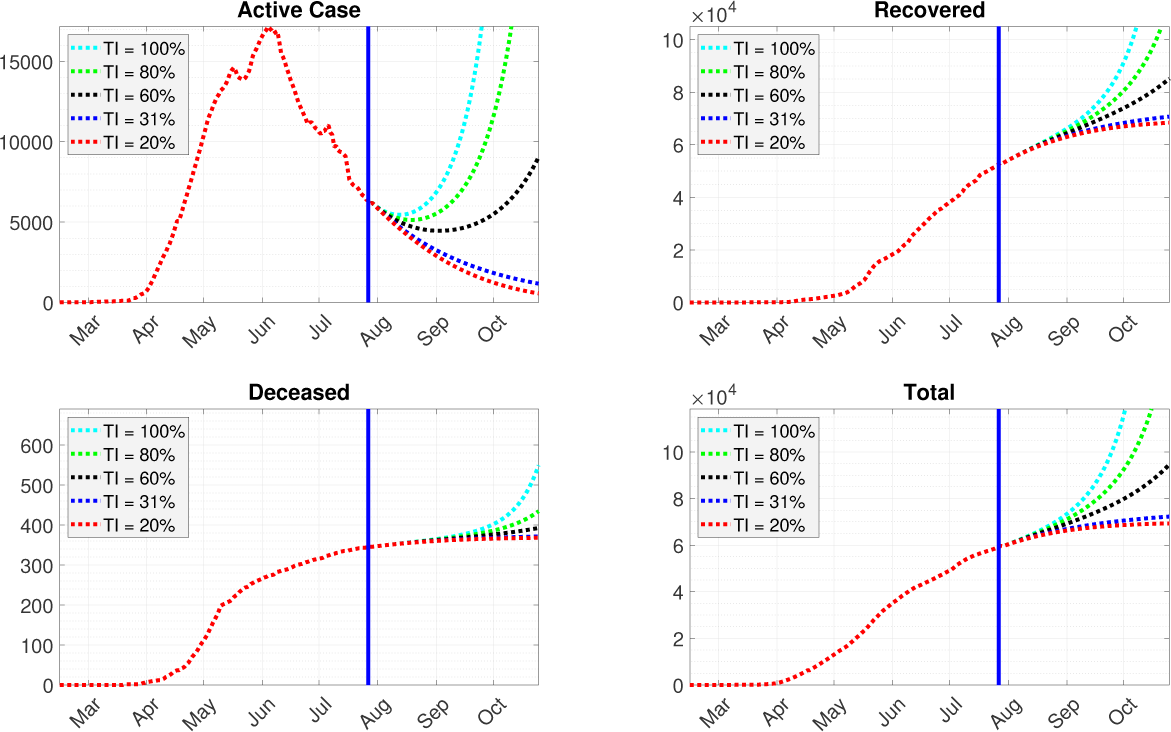
Forecasting for the next 90 days in UAE.

### 3.2. Australia

The first confirmed case of COVID-19 in Australia was found on January 2020 when a traveller was back to Victoria from Wuhan, China. The number of incidence passed 1000 on March 2020 and doubled after three days. The growth of incidence during March and April is considered as the first wave of pandemic with ℛ_*t*_ = 3 (see Figure 6). The effects of pandemic in health sectors started to reach other sectors such as trade, travel, economic and finance and an intensive interventions to prevent the pandemic from growing have been done [6].

**Figure 6:**
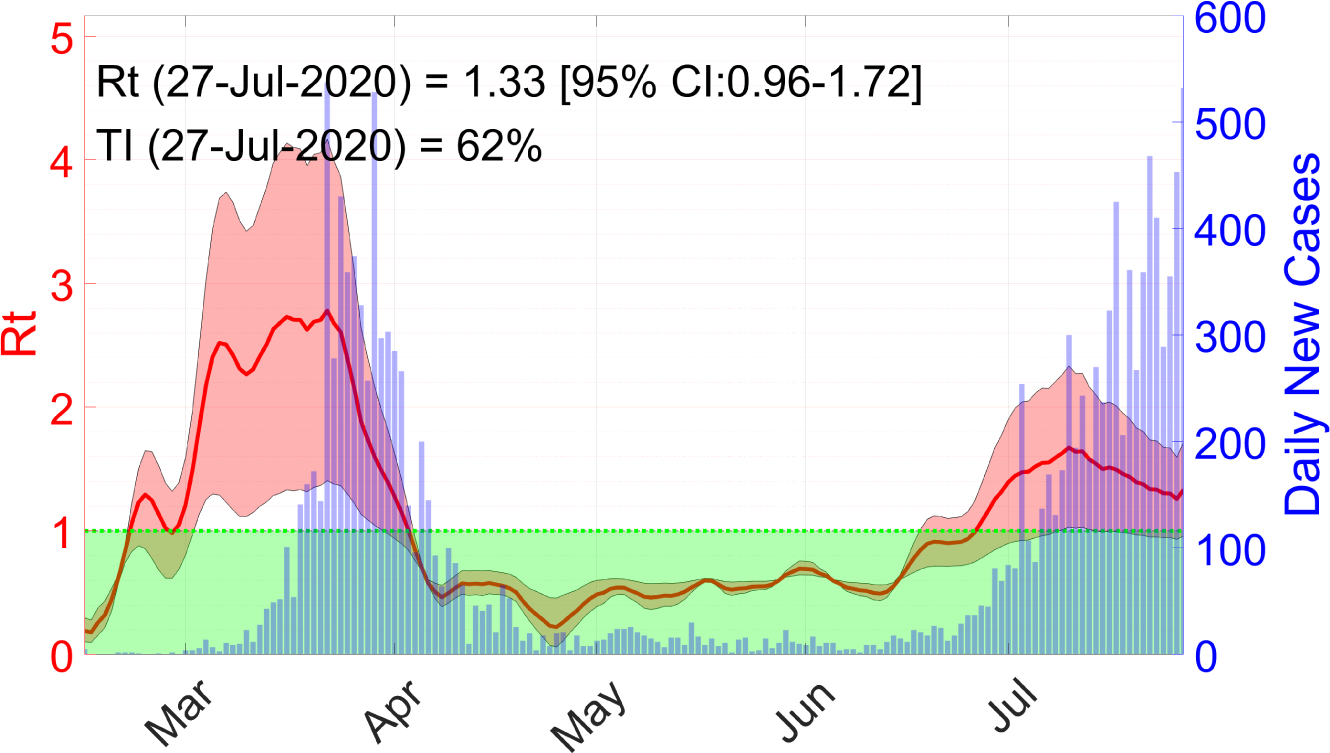
The time-varying effective reproduction number ℛ_*t*_ in Australia.

The Australian Government closed the borders to all non-residents and non-citizens on 20 March 2020 and applied a 14-day self isolation for all arrivals. Quarantine/lock-down related policy such as physical distancing or self-isolation policy has been applied in the form of school and workplace closure, mass gathering cancellation, contact tracing, etc. Also all the non-essential services have been stopped to maximize the physical distancing. The policy is applied for the next three months [24].

The effect of the interventions are indicated by the fall of ℛ_*t*_ *<* 1 in April, where the number of new cases dropped from 350 to 20 cases per day, as shown by Figure 6.

As the quarantine related policy is lifted in the beginning of June after a slow rate of infection (ℛ_*t*_ *<* 1) for three months, there has been a rise in the number of positive cases. This makes the Australian Government applies the policy softer in order to prevent not only higher cases in the second wave but also reducing a long-term impact to other than health sector. Australia TI by 27 July was 62% and its short-term projection shows that the active cases will increase sharply if there is no further interventions. The number of recovered and deceased cases will increase in the next 90. The short-term projection is shown in Figure 7.

**Figure 7:**
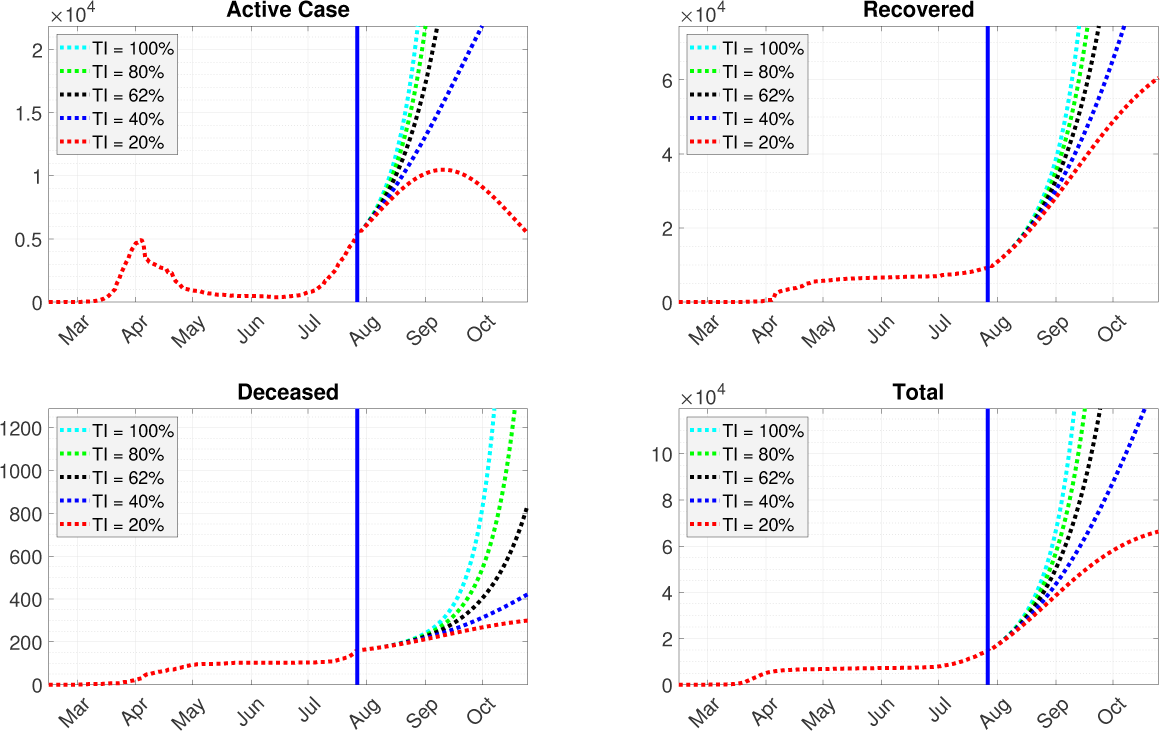
Forecasting for the next 90 days in Australia.

**Figure 8:**
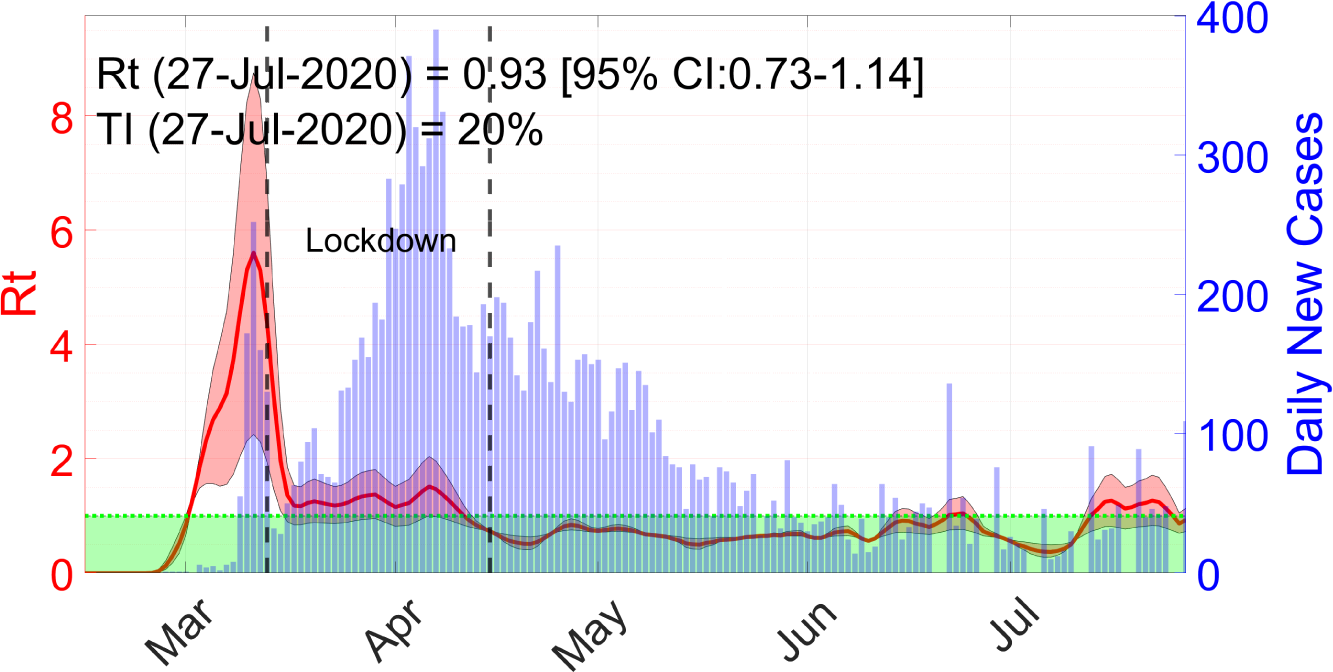
The time-varying effective reproduction number ℛ_*t*_ in Denmark.

**Figure 9:**
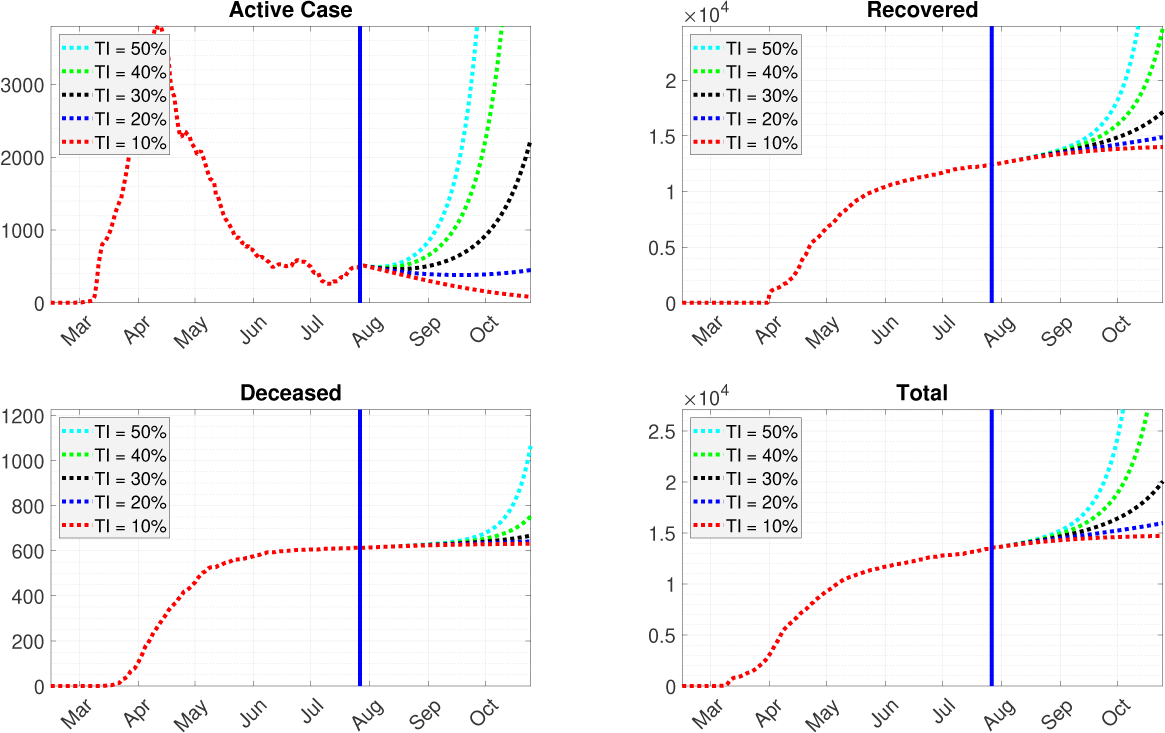
Forecasting for the next 90 days in Denmark.

### 3.3. Denmark

Denmark confirmed its first case on 27 of February, when a man who had been skiing in Lombardy in Italy returned to Denmark. Denmark introduced lock-down on 13 of March, by ordering people who are working in non-essential functions in the public sector to stay at home for two weeks. Furthermore, kindergarten, primary and secondary schools, universities, libraries, indoor cultural institutions and similar places were closed. Assemble more than ten people in public became illegal.

The effect of the lock-down can be observed after three weeks, when ℛ_*t*_ *<* 1. A very slow and gradual reopening has been initiated on 15 of April, by opening nurseries, kindergartens, and primary schools, However, the government will re-enforce lock-down if there are indications that the number of infections increases too fast. Universities are open only for employees, while all courses will be given online. At the end of July, there is an increase in the number of infected people, possibly due to the summer holiday. However, the government has not enforced any restriction.

Denmark TI by 27 July was 20%. A short-term projection shows the number of active cases will steady under the current measures. The number of recovered individuals will increase, while the number of death will decrease significantly.

## 4. Conclusion

We present a data-driven approach for modeling and forecasting of COVID-19 outbreak for public policy making. The method relies on the quality of the data. Thus, the the estimated ℛ_*t*_ and the forecast results need to be carefully interpreted when the number of testing is not sufficient. By estimating the Transmission Index (TI), our approach can be used for short to medium terms forecast based on policy to be taken by the decision makers. The forecast can be used to predict the course of COVID-19 including a probability of the upcoming second wave. Simulation results using data from three countries show our approach gives reasonable forecasts.

## Data Availability

Link to data and codes is written in the article.

https://github.com/agusisma/coviddatadriven

## References

[1] World Health Organization, WHO Coronavirus Disease (COVID-19) Dashboard, https://covid19.who.int (2020).

[2] U. Nation, Everyone included: Social impact of covid-19, https://www.un.org/development/desa/dspd/everyone-included-covid-19.html (2020).

[3] J. A. Lewnard, N. C. Lo, Scientific and ethical basis for social-distancing interventions against covid-19, The Lancet. Infectious diseases 20 (6) (2020) 631.

[4] J. Jia, J. Ding, S. Liu, G. Liao, J. Li, B. Duan, G. Wang, R. Zhang, Modeling the control of covid-19: Impact of policy interventions and meteorological factors, arXiv preprint 2003.02985.

[5] S. H. Khoshnaw, M. Shahzad, M. Ali, F. Sultan, A quantitative and qualitative analysis of the covid–19 pandemic model, Chaos, Solitons & Fractals (2020) 109932.

[6] S. L. Chang, N. Harding, C. Zachreson, O. M. Cliff, M. Prokopenko, Modelling transmission and control of the covid-19 pandemic in australia, arXiv preprint 2003.10218.

[7] L. Pang, S. Liu, X. Zhang, T. Tian, Z. Zhao, Transmission dynamics and control strategies of covid-19 in wuhan, china, Journal of Biological Systems (2020) 1–18.

[8] G. Grasselli, A. Pesenti, M. Cecconi, Critical care utilization for the covid-19 outbreak in lombardy, italy: Early experience and forecast during an emergency response, JAMA 323 (2020) 1545–1546. doi:10.1001/jama.2020.4031.

[9] A. Hasan, H. Susanto, V. R. Tjahjono, R. Kusdiantara, E. R. M. Pu- tri, P. Hadisoemarto, N. Nuraini, A new estimation method for covid-19 time-varying reproduction number using active cases (2020). 2006.03766.

[10] M. Egger, L. Johnson, C. Althaus, A. Schöni, G. Salanti, N. Low, S. L. Norris, Developing who guidelines: time to formally include evidence from mathematical modelling studies, F1000Research 6.

[11] H. Susanto, V. Tjahjono, A. Hasan, M. Kasim, N. Nuraini, E. Putri, R. Kusdiantara, H. Kurniawan, How many can you infect? simple (and naive) methods of estimating the reproduction number, arXiv preprint 2006.15706.

[12] C. Anastassopoulou, L. Russo, A. Tsakris, C. Siettos, Data-based analysis, modelling and forecasting of the covid-19 outbreak, PLoS ONE 15. doi:10.1371/journal.pone.0230405.

[13] K. Roosa, Y. Lee, R. Luo, A. Kirpich, R. Rothenberg, J. Hyman, P. Yan, G. Chowell, Real-time forecasts of the covid-19 epidemic in china from february 5th to february 24th, 2020, Infectious Disease Modelling 5 (2020) 256–263. doi:10.1016/j.idm.2020.02.002.

[14] D. Fanelli, F. Piazza, Analysis and forecast of covid-19 spreading in china, italy and france, Chaos, Solitons & Fractals 134. doi:10.1016/j.chaos.2020.109761.

[15] K. K.-W. To, O. T.-Y. Tsang, W.-S. Leung, A. R. Tam, T.-C. Wu, D. C. Lung, C. C.-Y. Yip, J.-P. Cai, J. M.-C. Chan, T. S.-H. Chik, D. P.-L. Lau, C. Y.-C. Choi, L.-L. Chen, W.-M. Chan, K.-H. Chan, J. D. Ip, A. C.-K. Ng, R. W.-S. Poon, C.-T. Luo, V. C.-C. Cheng, J. F.-W. Chan, I. F.-N. Hung, Z. Chen, H. Chen, K.-Y. Yuen, Temporal profiles of viral load in posterior oropharyngeal saliva samples and serum antibody responses during infection by sars-cov-2: an observational cohort study, The Lancet Infectious Diseases 20 (2020) 565–74. doi:10.1016/S1473-3099(20)30196-1.

[16] O. W. in Data, Coronavirus (covid-19) testing, https://ourworldindata.org/coronavirus-testing (2020).

[17] N. Golding, T. W. Russell, S. Abbott, J. Hellewell, C. A. B. Pearson, K. van Zandvoort, C. I. Jarvis, H. Gibbs, Y. Liu, R. M. Eggo, J. W. Edmunds, A. J. Kucharski, Reconstructing the global dynamics of under-ascertained covid-19 cases and infections (2020). 2020.07.07.20148460.

[18] K. Nandkeolyar, Coronavirus in uae: Four of a family infected, Gulf News. Archived from the original on 29 January 2020. Retrieved 29 January 2020. (2020).

[19] G. Duncan, S. Gautam, Coronavirus: Uae records first case, The National. Archived from the original on 29 January 2020. Retrieved 29 January 2020. (2020).

[20] Why uae school closures are an important opportunity to fight coronavirus, The National. Retrieved 13 April 2020. (2020).

[21] Coronavirus: Prayers at mosques and all other places of worship in uae suspended, gulfnews.com. Retrieved 13 April 2020. (2020).

[22] Coronavirus: Uae imposes night curfew as it carries out disinfection campaign, Middle East Eye. Retrieved 26 March 2020. (2020).

[23] N. E. Crisis, D. M. Authority, Uae coronavirus (covid-19) updates, may 18, 2020, https://covid19.ncema.gov.ae/en/News/Details/1270 (2020).

[24] K. Moloney, S. Moloney, Australian quarantine policy: From centralization to coordination with mid-pandemic covid-19 shifts, Public Administration Review 80 (4) (2020) 671–682. doi:10.1111/puar.13224.

